# Effect of one-size-fits-all amplification in Bluetooth hearing devices for hearing impaired listeners’ speech recognition

**DOI:** 10.1101/2023.03.08.23287011

**Authors:** Neila Bell, Leah Gibbs, Jusung Ham, Kayla Howerton, Inyong Choi, Jaeseong Lee, Kyoung Ho Bang, Han-gil Moon

## Abstract

Hearing loss is a highly prevalent chronic condition that degrades the quality of life. Although hearing aids provide immediate and efficient benefits to listeners with mild-to-moderate hearing loss, the prevalence of hearing aid use has been low. Consumer wireless earbuds are increasingly being equipped with the ability to amplify external sounds, which can be an affordable alternative to hearing aids. This study compared the amplification performance of non-customized Bluetooth consumer hearables to high-end hearing aids when used by people with mild-to-moderate hearing loss. We found that such a non-customized consumer device significantly enhances the speech recognition of listeners with mild-to-moderate hearing loss, although its performance did not reach the hearing aids. These results determine the extent to which inexpensive and accessible non-customized Bluetooth hearables can help people with mild-to-moderate hearing loss.

## 1. Introduction

Hearing loss is one of the most common chronic conditions in the United States (Blackwell et al., 2014). Its prevalence is expected to increase as life expectancy increases; the prevalence of hearing loss in the United States is expected to double by 2060 (Goman et al., 2017). Hearing loss limits communication, which causes emotional distress, social isolation, and reduced quality of life (Shukla et al., 2020, 2021). Furthermore, hearing loss has been identified as a “potentially modifiable” risk factor for dementia in midlife (Livingston et al., 2020), making early treatment of hearing loss critical.

The most immediate and effective treatment for hearing loss is the use of hearing aids (Shukla et al., 2021). However, 75% of people with hearing loss do not use hearing aids (Chien & Lin, 2012). The main barriers to hearing aid use appear to be the high cost, complex and time-consuming acquisition process, and social stigma (Reed et al., 2019).

Personal Sound Amplification Products (PSAPs) are consumer electronic devices that amplify sounds. These devices can be purchased and fitted by the user themselves and are much less expensive than hearing aids. While not classified as a medical device that can “cure” hearing loss, recent studies have shown that PSAPs offer similar benefits to hearing aids in terms of speech perception and listening effort (Chen et al., 2022; Mamo et al., 2016; Manchaiah et al., 2017; Reed et al., 2017). However, there is considerable variation in performance among the many PSAP products (Brody et al., 2018), making it necessary to investigate the performance of specific popular PSAPs.

Consumer wireless earbuds, or true wireless stereo (TWS) hearables, are increasingly being equipped with the ability to amplify external sounds, which can be a very affordable alternative to hearing aids. A recent study found that Apple’s AirPods Pro, which have the largest share of the TWS market, performed comparably to hearing aids (Lin et al., 2022). The AirPods Pro work with Apple’s iPhone products to provide external sound amplification tailored to the user’s hearing by inputting an individual’s audiogram.

On the other hand, Samsung’s Galaxy Buds 2 Pro, which has the second largest share of the TWS market, provides external sound amplification for the most common types of mild-to-moderate hearing loss without requiring the user to enter their audiogram. The purpose of this study is to compare the amplification performance of these non-customized Bluetooth TWS hearables when used by people with mild-to-moderate hearing loss, and the extent to which they improve users’ speech perception, with unaided conditions and high-end hearing aids. In doing so, we hope to determine the extent to which inexpensive and accessible non-customized Bluetooth TWS hearables can help people with mild-to-moderate hearing loss.

## 2. Material and Methods

### 2.1 Participants

A total of 30 adults, 20 female 10 male, ages 55 to 86 years old, were enrolled as participants. Participants had a range of experience with hearing assistive devices including using hearing aids (HA), over the counter (OTC) devices, and having no experience at all. Participants had bilateral mild-to moderate hearing loss.

All participants provided written informed consent. Participants were tested at the Hearing Aid and Research Laboratory at the University of Iowa. The protocol followed by the study was approved by the IRB no. Subjects were compensated at an hourly base rate of $20/hr., and received parking vouchers for the duration of their visit.

### 2.2 Devices

Three conditions were tested: 1) unaided, 2) Samsung Galaxy Buds2 Pro (henceforth referred to as “GB2P”), and 3) Oticon More 2 Has (“HA”).

If the patient’s audiometric thresholds were within 6 months, a repeat evaluation was not needed, and devices were programmed with their threshold data before their visit. If past 6 months, air conduction thresholds were obtained first, and then used for device programming. Oticon More 2 hearing aids were receiver-in-canal style and were programed to manufacturer first fit with appropriate dome selection and full gain adaptation. No other changes to the software were made, and new firmware updates were not installed post pilot participant which most closely mirrored the “first fit” function of the other two wearables. Samsung Galaxy wearables (GB2P) were in “Amplify Ambient Sound” mode for all tasks.

### 2.3 “Real-Ear” Measures

First, otoscopy was performed to check for clear healthy canals. Real ear aided response (REAR) measures were completed for all 4 testing conditions using Audioscan Verifit®2. Equipment was calibrated before every participant, and probe tube depth within 5mm of the eardrum was verified using the probe tube placement tool ProbeGUIDE™. All available air conductio audiometric data was entered, the NAL-NL2 formula was chosen for targets, and stimulus for all conditions was an average speech sample. Data was collected to evaluate root mean square error (RMSE), speech intelligibility index (SII), and frequency response.

### 2.2 Task design and procedures

Patients were asked to perform three speech understanding tasks. Two were the Hearing In Noise Test (HINT) given in noise and in quiet, in a randomized order, to establish patients sentence speech reception thresholds (sSRTs). The stimuli constituted of 25 phonemically balanced lists (Nilsson et al. 1994) also in a randomized order for every participant. For the noise condition, the stimuli and masker were delivered through one loudspeaker (spatially coincident) that the participant was coached to face directly. The masker was HINT noise at 70dB HL for all conditions and participants.

Iowa Test of Consonant Perception [ITCP: (Geller et al., 2021)]: In a custom-written Matlab script, participants were presented with a crosshair on a screen and were instructed to listen while a word was played auditorily. The following screen consisted of four words; the target word and three other words differing by a phoneme. The participant was instructed to select the word they heard via a keyboard. Within a single condition, there were 120 trials with built-in voluntary breaks every 30 trials. Stimulus level was decided via a test trail of 10 trials starting at 57dB HL. If the participant scored 50% or better, all conditions ITCP was set at 57dB. If <50%, the test trials were repeated by increasing stimuli intensity by 3 dB until 50% was achieved. These three tests were repeated for each condition, with a break in the middle to help prevent participant fatigue.

## 3. Results

### 3.1 Real Ear Measures

Figure 1 shows REAR-measured frequency-response curves averaged across all of the 30 subjects for each of unaided, GB2P, and HA conditions compared to the NAL-NL2-based target curve. Both GB2P and HA show significantly higher gains across all the tested frequencies than the unaided condition. GB2P shows significantly lesser gain than HA above 2,000Hz.

**Figure 1.**
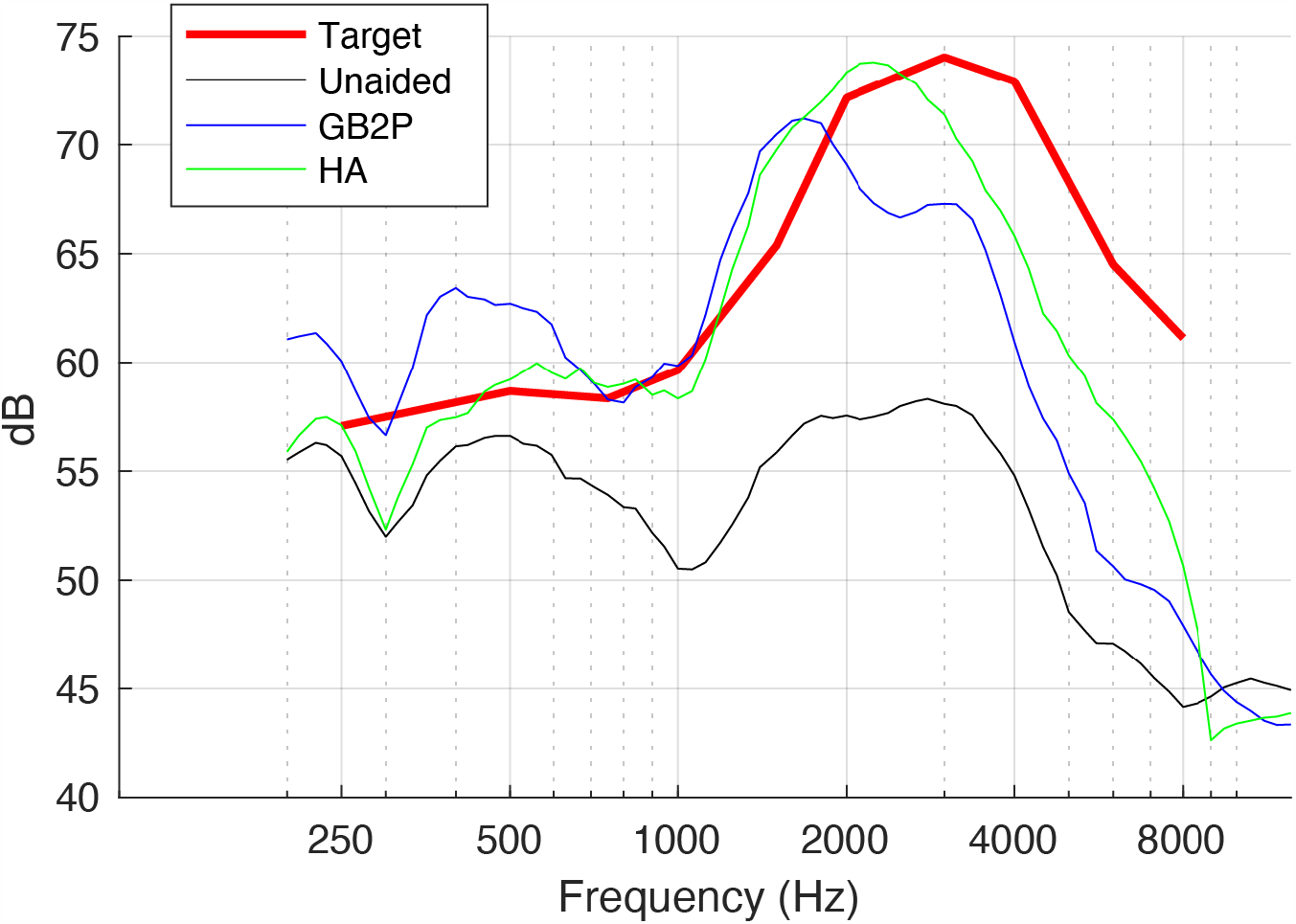
Real ear aided response (REAR)-measured frequency-response curves averaged across all of the 30 subjects.

### 3.2 Speech Intelligibility Index

Figure 2 shows pairwise comparisons of Speech Intelligibility Index (SII). For both left and right ears, both GB2P and HA show significantly higher SII than the unaided condition. When GB2P and HA were compared, HA exhibited significantly higher SII than the GB2P.

**Figure 2.**
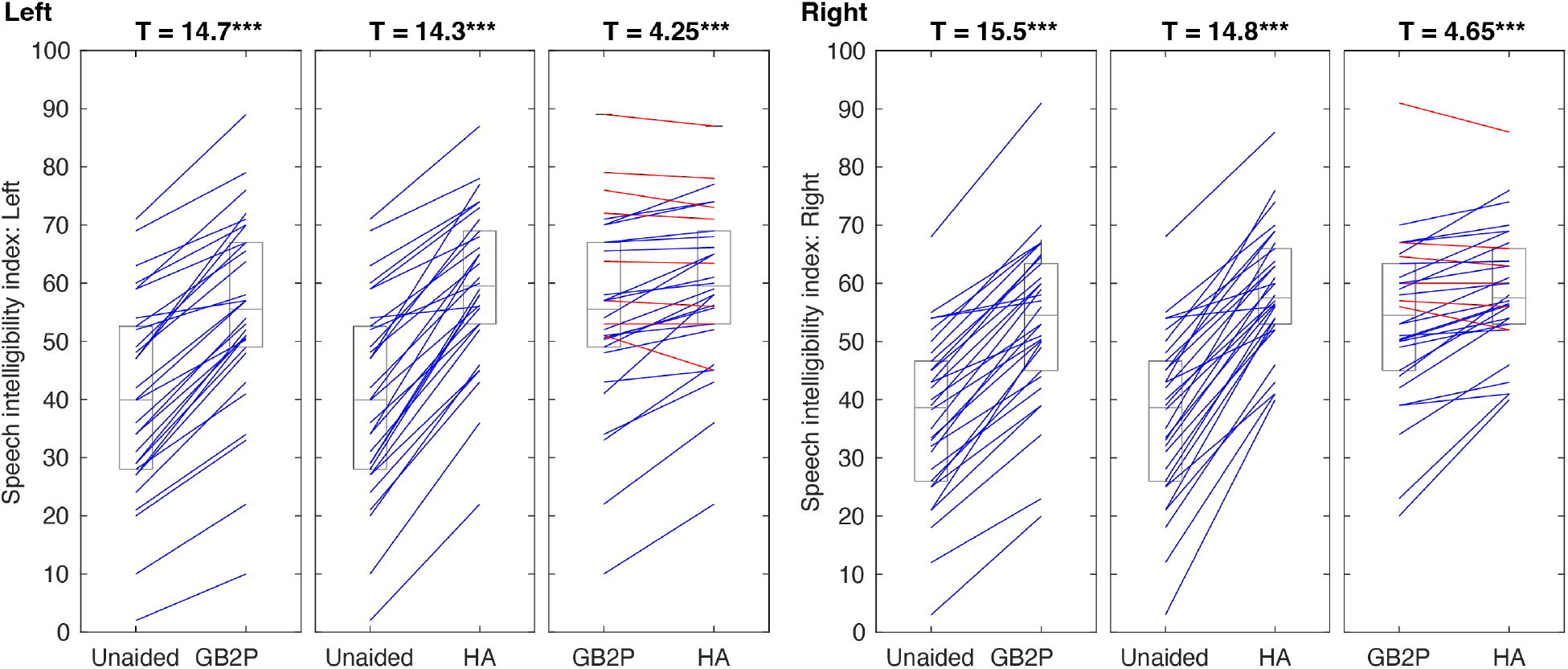
Pairwise comparisons of Speech Intelligibility Index (SII). Three asterisks (***) indicate a statistically significant difference between conditions at the p<0.001 level (paired t-test).

### 3.2 Speech recognition performance

Figure 3 shows pairwise comparisons of speech perception performance. For single word recognition, both GB2P and HA show significantly higher accuracy (percent correct) than the unaided condition. When GB2P and HA were compared, HA exhibited significantly higher performance than the GB2P. For sentence recognition, both GB2P and HA show significantly lower speech-reception thresholds (SRTs) than the unaided condition. When GB2P and HA were compared, HA exhibited significantly lower SRTs than the GB2P.

**Figure 3.**
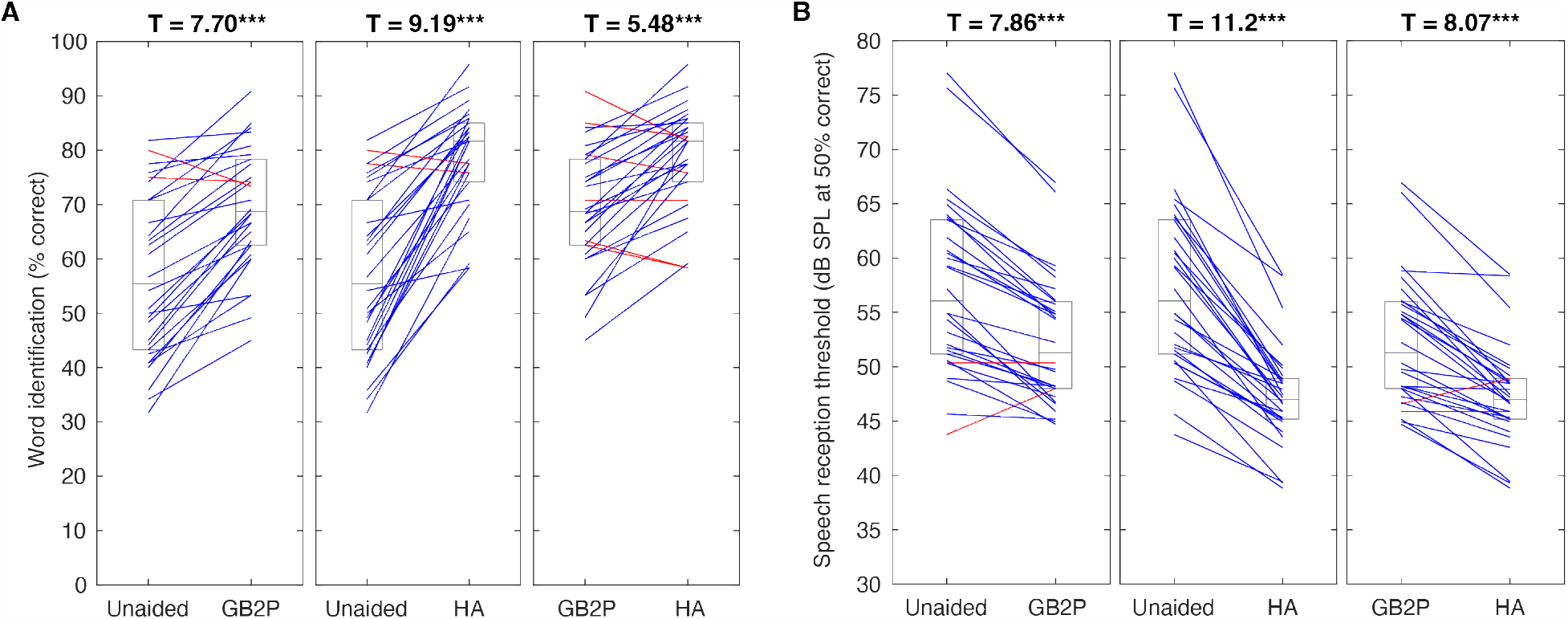
A Pairwise comparisons of single word-identification performance. B Pairwise comparisons of speech reception thresholds (dB SPL at 50% correct). Three asterisks (***) indicate a statistically significant difference between conditions at the p<0.001 level (paired t-test).

### 3.2 Relationship between audiometry and the amplification benefit

Figure 4 shows relationships between left-ear 3kHz pure-tone thresholds and the amplification benefit in SRTs (i.e., unaided - aided). Pearson correlation coefficients were 0.607 (p<0.001) for HA and 0.360 (p = 0.051, uncorrected) for GB2P.

**Figure 4.**
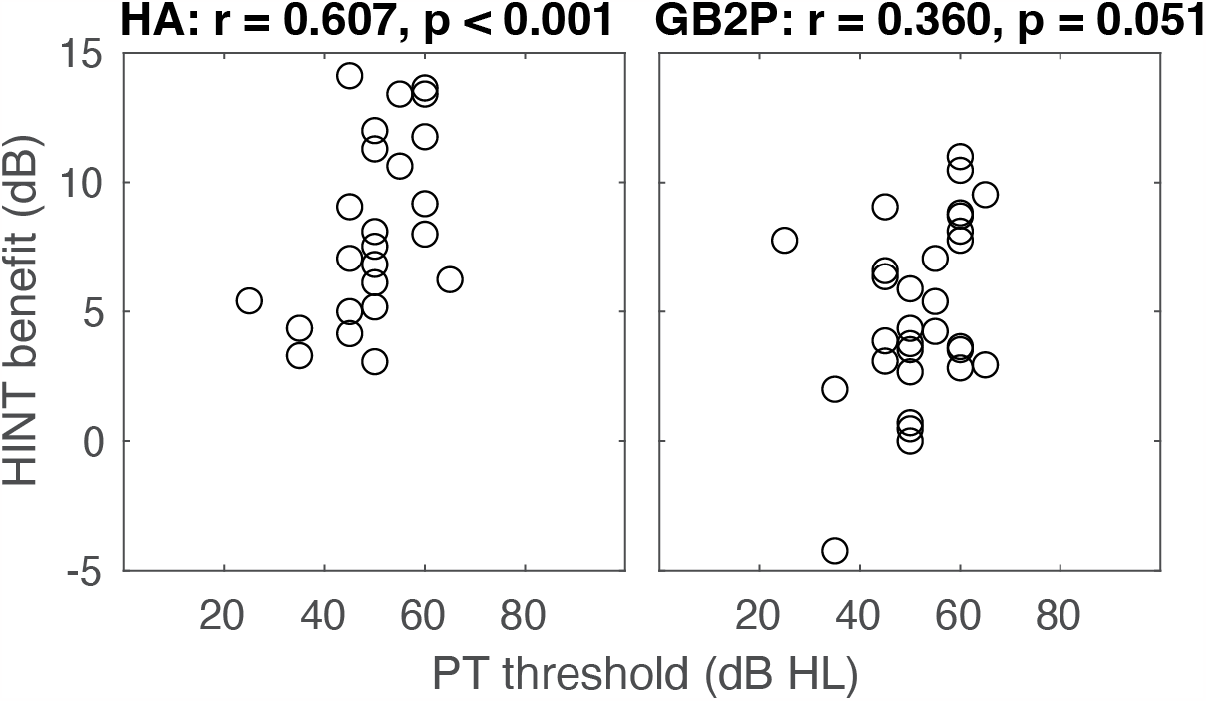
Relationships between left-ear 3kHz pure-tone thresholds and the amplification benefit in speech reception thresholds (i.e., unaided – aided). Left panel: HA, Right panel: GB2P. r values are Pearson correlation coefficients.

## 4. Discussion

The purpose of this study was to observe how much the non-customized amplification function of smartphone-bundled earphones improve speech perception in listeners with symmetrical mild-to-moderate hearing loss. Using Samsung’s Galaxy Buds2 Pro in the “Ambient Sound” mode with the “Amplify Ambient Sound” feature, most patients with mild to moderate hearing loss showed improved speech perception compared to not using the device. Although the performance was modest compared to the high-performance hearing aids we compared it to, these results suggest that the selected product (Samsung’s Galaxy Buds2 Pro) has the potential to serve as an appropriate hearing aid-experience for people with mild to moderate hearing loss.

In the presence of background noise, the high-performance hearing aids showed a significant improvement in speech recognition, while the Galaxy Buds2 Pro did not.

Although not classified as a “medical device” or designed to treat the hearing-impaired population based on the US Food and Drug Administration’s (FDA) draft guidance, PSAPs are thought to compensate for hearing loss. PSAPs, including the Galaxy Buds2 Pro tested in this study, offer a significantly lower cost and greater accessibility than traditional hearing aids, making them a good gateway to the hearing aid experience for people with hearing loss who have not yet experienced it.

PSAPs, such as Apple’s AirPods and Samsung’s Galaxy Buds tested in this study, are considered socially trendy. Given that social stigma is one of the reasons many people with hearing loss do not use hearing aids, the design of these user devices can be a positive factor in the hearing aid experience.

While the specific PSAPs tested in this study do improve speech perception for people with mild to moderate hearing loss, several issues need to be addressed for better performance. First, customized amplification needs to be provided. Unlike high-performance hearing aids, Galaxy Buds, which provided the same amount of amplification for all the subjects, did not tend to provide greater speech perception gains with more severe hearing loss. This is likely due to the lack of customizable amplification. While one of the main selling points of PSAPs is the convenience of “no fitting process,” the inability to customize to the user’s hearing can lead to a poor impression of PSAPs by people with hearing loss.

It should also be able to improve speech recognition in noisy environments. It would be desirable to include noise reduction features commonly found in hearing aids, microphones that amplify only the sound in front of them, or signal processing algorithms that distinguish between background noise and the sounds that need to be heard.

## 5. Conclusion

In conclusion, our study showed that the ambient sound amplification feature of Samsung Galaxy Buds2 Pro, a wireless earphone bundled with a smartphone, significantly improved speech perception in patients with mild to moderate hearing loss. With future improvements and quality control, these devices could be an excellent means of bringing the hearing aid experience to people with hearing loss who have not yet experienced it, at a lower cost and with less social stigma.

## Data Availability

All data produced in the present study are available upon reasonable request to the authors.

## Acknowledgments

The authors declare no competing financial interests.

